# A quantitative evaluation of aerosol generation from upper airway suctioning during tracheal intubation and extubation sequences

**DOI:** 10.1101/2021.12.12.21267658

**Authors:** AJ Shrimpton, JM Brown, TM Cook, CM Penfold, JP Reid, AEP Pickering

## Abstract

**Background:** Open respiratory suctioning is considered to be an aerosol generating procedure (AGP) and laryngopharyngeal suction, used to clear secretions during anaesthesia, is widely managed as an AGP. It is uncertain whether such upper airway suctioning should be designated an aerosol generating procedure (AGP) because of a lack of both aerosol and epidemiological evidence of risk.

**Aim:** To assess the relative risk of aerosol generation by upper airway suction during tracheal intubation and extubation in anaesthetised patients.

**Methods:** Prospective environmental monitoring study in ultraclean operating theatres to assay aerosol concentration during intubation and extubation sequences including upper airway suctioning for patients undergoing surgery (n=19 patients). An Optical Particle Sizer (particle size 300nm-10μm) was used to sample aerosol from 20cm above the patient’s mouth. Baseline recordings (background, tidal breathing and volitional coughs) were followed by intravenous induction of anaesthesia with neuromuscular blockade. Four periods of oropharyngeal suction were performed with a Yankauer sucker: pre-laryngoscopy, post-intubation and pre- and post-extubation.

**Findings:** Aerosol from breathing was reliably detected (65[39-259] particles.L^−1^ (median[IQR])) above background (4.8[1-7] particles.L^−1^, p<0.0001 Friedman). The procedure of upper airway suction was associated with much lower average concentrations of aerosol than breathing (6.0[0-12] particles.L^−1^, P=0.0007) and was indistinguishable from background (P>0.99). The peak aerosol concentration recorded during suctioning (45[30-75] particles.L^−1^) was much lower than both volitional coughs (1520[600-4363] particles.L^−1^, p<0.0001, Friedman) and tidal breathing (540[300-1826] particles.L^−1^, p<0.0001, Friedman).

**Conclusion:** The procedure of upper airway suction during airway management is associated with no higher concentration of aerosol than background and much lower than breathing and coughing. Upper airway suction should not be designated as a high risk AGP.

## Background

Respiratory tract suctioning and open suctioning of the airways are classified as aerosol generating procedures (AGPs) [1,2]. This designation is based on weak epidemiological data suggesting an increased risk of transmission during the SARS 2003 outbreak [3]. The Public Health England AGP guidelines state *“the consensus view of the UK IPC cell is that only open suctioning beyond the oro-pharynx is currently considered an AGP”* [2]. A recent systematic review concluded it was “*not possible to establish a clear absence of risk associated with [airway suctioning]”* due to the lack of evidence [4]. Specifically, there are no clinical aerosol studies to help inform any risk assessments for airway suction.

Previous clinical aerosol monitoring studies during tracheal intubation and extubation, facemask ventilation and supraglottic airway use demonstrated these airway management procedures generate less aerosol than a patient’s own natural respiratory activities [5–7]. Placement of a Yankaeur-type sucker into the laryngopharynx to clear secretions around the glottic inlet prior to tracheal intubation and following extubation constitutes open-suctioning beyond the oropharynx. As such, this intervention commonly performed during tracheal intubation / extubation sequences, is being managed as an AGP which necessitates the use of airborne personal protective equipment (PPE) and fallow time after its performance. To address the question of whether upper airway suction is an AGP we undertook clinical aerosol monitoring during tracheal intubation and extubation sequences with pre-specified periods of upper airway suction in patients undergoing general anaesthesia for surgery. Aerosol generated during upper airway suction was compared against the patient’s own coughs and breathing to assess the relative risk associated with the procedure.

## Methods

This study was performed as part of the NIHR funded MAGPIE study (within the AERATOR group of studies). Ethical approval was granted by the Greater Manchester REC committee (Reference: 20/NW/0393, approved 18/09/2020) and all patients gave written informed consent. The study was granted Urgent Public Health status by NIHR and is registered in the ISRCTN registry (ISRCT:N21447815).

We undertook a prospective environmental monitoring study in a UK hospital (Southmead Hospital, North Bristol NHS Trust). All recordings were undertaken in operating theatres with ultraclean ventilation systems (EXFLOW 32, Howorth Air Technology, Farnworth, UK) placed in standby mode. We have previously described in detail the effect of placing this ventilation system in standby mode [8,9]. This results in an ultraclean environment with an air exchange rate equivalent to standard UK operating theatres.

Aerosol monitoring was performed using an optical particle sizer (OPS, TSI Incorporated, Model 3330, USA) which detects particles from 300 nm to 10 μm diameter, reporting the recorded size distribution, number and mass concentrations over a 1 s interval. A 3D printed funnel (90 mm height, maximum diameter of 75 mm and 10 mm exit port, formed of polylactic acid on a RAISE3D Pro2 Printer, 3DGBIRE, UK) was attached to the OPS via a 1.25 m length of conductive silicone tubing (TSI Incorporated, Model 3330, USA).

All patients were over 18 years of age and were scheduled for either elective (n=11) or emergency (n=8) surgery. The elective patients were on low risk ‘Green’ pathways with respect to COVID-19 (i.e. asymptomatic, negative COVID-19 polymerase chain reaction (PCR) test in the previous 72 hours and had self-isolated since this test). Patients undergoing emergency surgery, were asymptomatic for COVID-19 but had not self-isolated prior to admission and either had a negative COVID-19 PCR test result or had a PCR test pending – these patients were treated as having indeterminate risk for COVID-19 (‘Amber’ pathway).

All participants were positioned supine with head positioning and airway management as per the anaesthetist’s preference. The research team were not involved in the provision of anaesthetic care. All members of the research and anaesthetic team wore personal protective equipment according to hospital protocol depending on the COVID-19 risk status of the patient.

Aerosol measurements of particle number concentration were performed with the sampling funnel positioned 20 cm above the patient’s mouth. A standardised protocol was followed for each patient:

Baseline aerosol sampling (performed before induction of anaesthesia) which comprised measuring background (with the funnel directed away from the patient), 60 seconds of tidal breathing and three volitional coughs.

After pre-oxygenation with 100% oxygen, anaesthesia was induced with intravenous propofol plus an opioid followed by neuromuscular blockade. Four phases of upper airway suction were specified:

Pre-intubation: immediately before laryngoscopy (after neuromuscular blockade),

Post-intubation: immediately after tracheal intubation (after cuff inflation),

Pre-extubation: before tracheal extubation (before cuff deflation) and

Post-extubation: after tracheal extubation (if tolerated by the patient).

All periods of upper airway suctioning were performed for a minimum of 10 seconds using a Yankauer suction device (with the side-port occluded) connected to an active negative pressure suction cannister as part of the anaesthetic machine (Aisys CS2, GE Healthcare, Chicago, IL, USA) set to max (−35kPa). Additional suctioning was undertaken as clinically indicated.

Sampling was continuous during the whole intubation sequence from baseline breathing measures until the second period of upper airway suctioning following tracheal intubation (Figure 1). Aerosol sampling was recommenced prior to tracheal extubation to include both periods of upper airway suctioning.

**Figure 1.**
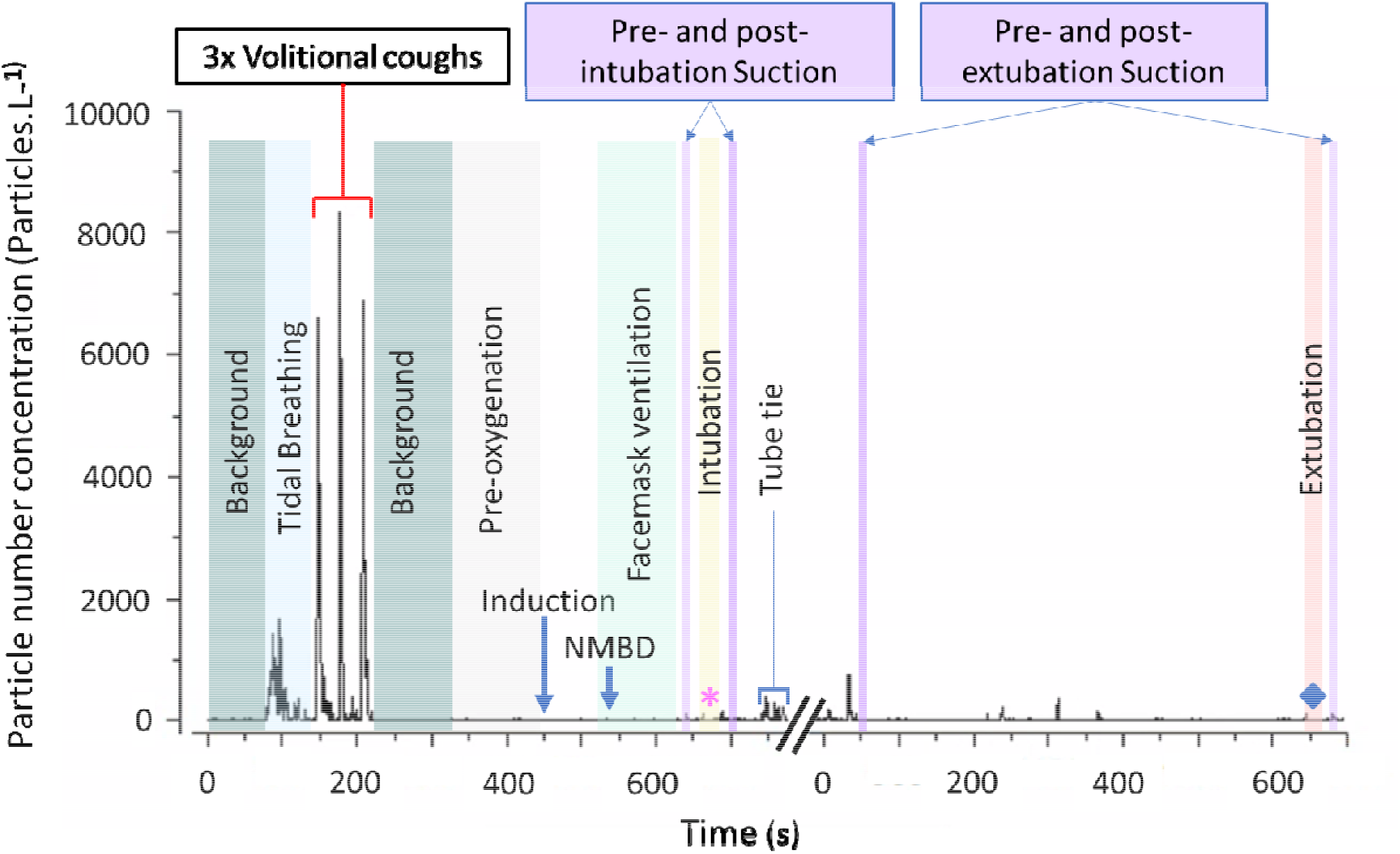
Timeline of aerosol concentration associated with upper airway suction during the intubation and extubation sequences in a representative patient. This shows the number concentration of particles detected during baseline respiratory manoeuvres (tidal breathing and voluntary coughs), background monitoring, facemask ventilation, intubation (from laryngoscopy until inflation of tracheal tube cuff), extubation (tracheal tube cuff down until tube out) and phases of suction. Note break in recording shown on x-axis between intubation and extubation sequences (* = tracheal intubation, ♦ = tracheal extubation). NMBD = neuromuscular blocking drug. Particle size range 300nm-10μm.

All events were timestamped by the researcher, these included: baseline tidal breathing, coughs, induction of anaesthesia, neuromuscular blockade administration, facemask ventilation, airway adjunct insertion, each period of airway suctioning, laryngoscopy, tracheal intubation, tracheal cuff inflation, tracheal cuff deflation and tracheal extubation.

It is currently unknown what aerosol emission rate is clinically significant with regard to the risk of respiratory pathogen transmission, and how this is reflected in the number concentration of aerosol at the source. In our facemask ventilation study, the mean (SD) particle concentration for tidal breathing was 303 (375) particles.L^−1^. We considered a 2-fold increase in the average aerosol concentration above breathing to be potentially meaningful (albeit still considerably lower than that associated with a cough). A priori sample size calculations for a non-parametric paired comparison predicted that recordings of complete data on 12 participants would ensure the study was adequately powered to detect a difference of this magnitude when compared to the patient’s own tidal breathing (80% power, alpha 0.05, G*Power3.1.9.4).

The TSI Aerosol Instrument Manager software was used to process the aerosol data prior to analysis in Origin Pro (Originlab, Northampton, Massachusetts, USA) and R (Version 4.1.2). The Shapiro-Wilk test was used to assess the normality of data. Comparisons were made between aerosol measurements using a repeated measures analysis for non-parametric data (Friedman with Dunn’s multiple comparisons with Bonferroni correction). Separate analyses were performed for the intubation sequences (n=18), extubation sequences (n=13) and peak values (n=19) to account for missing values. The significance level was set at P < 0.05. All data are presented as mean (SD) or median (IQR).

## Results

The aerosol monitoring study was performed over a 7-week period during anaesthesia for patients having orthopaedic, neurosurgical and general surgical procedures. Following informed consent 19 patients were recruited (female=9, age 70 (10) yrs, BMI 28.7 (5.6) kg.m_-2_, mean (SD)).

The airway of each patient was managed with a cuffed oral tracheal tube. During facemask ventilation (before tracheal intubation) four patients had a Guedel oropharyngeal airway inserted to facilitate ventilation. Neuromuscular blockade was provided with rocuronium (n=18) or atracurium (n=1).

All patients (n=19) underwent pre-intubation and post-intubation upper airway suction. Fourteen patients underwent pre-extubation upper airway suction and all but one of these tolerated post-extubation upper airway suction. Therefore, during the conduct of the study, we recorded aerosol generation through 19 entire intubation sequences and 14 extubation sequences (including periods of facemask ventilation). A total of 784 seconds of aerosol data were recorded during episodes of suction (410 s peri-intubation, 374 s peri-extubation).

In eight instances tracheal tube placement required the use of an adjunct: a bougie in seven and a stylet in one. One patient had a rapid sequence induction of anaesthesia for emergency surgery and no period of facemask ventilation but did have pre-intubation suction. In one patient an accidental oesophageal intubation was identified following cuff inflation and absence of CO_2_ noted on capnography. This patient had the tracheal tube removed, underwent a further period of facemask ventilation and was successfully intubated at the second attempt with the assistance of a bougie but without any further suction. Both intubation attempts are included in the analysis and post-intubation suction was performed after the airway had been secured. One patient coughed twice following tracheal extubation: once immediately after extubation and once during post-extubation suctioning.

Background aerosol concentrations were (median[IQR]) 4.8 [1.0-6.8] particles.L^−1^). Aerosol generated by tidal breathing was reliably detected above background levels (65 [39-259] particles.L^−1^), p<0.0001, Friedman) (Figures 1–3). Aerosol concentrations detected during periods of facemask ventilation, pre-intubation suction, tracheal intubation (from laryngoscopy until cuff up) and post-intubation suction were not statistically different from background levels (all p>0.99) or each other (Table 1 and Figure 2). The concentration of aerosol detected during suctioning pre- and post-tracheal intubation and pre-tracheal extubation were far lower than tidal breathing (all p<0.0001, Friedman) (See Table 1 and Figure 3).

**Table 1.**
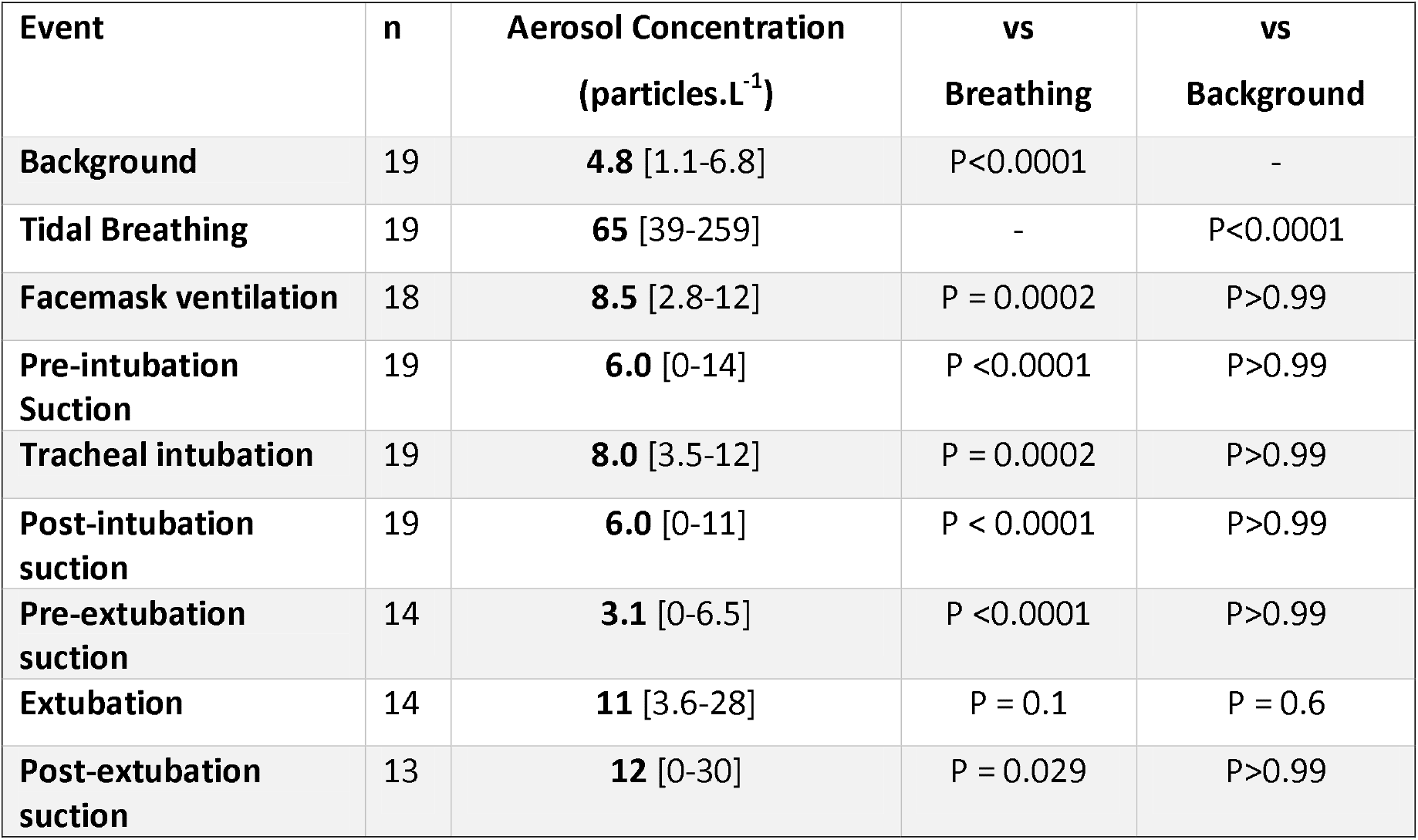
Aerosol concentrations measured during different procedures compared to tidal breathing and background (Friedman with Dunn’s multiple comparisons). (median [IQR])

| **Event** | **n** | **Aerosol Concentration**<br>**(particles.L<sup>-1</sup>)** | **vs**<br>**Breathing** | **vs**<br>**Background** |
| --- | --- | --- | --- | --- |
| **Background** | 19 | **4.8** [1.1-6.8] | P<0.0001 | - |
| **Tidal Breathing** | 19 | **65** [39-259] | - | P<0.0001 |
| **Facemask ventilation** | 18 | **8.5** [2.8-12] | P = 0.0002 | P>0.99 |
| **Pre-intubation**<br>**Suction** | 19 | **6.0** [0-14] | P <0.0001 | P>0.99 |
| **Tracheal intubation** | 19 | **8.0** [3.5-12] | P = 0.0002 | P>0.99 |
| **Post-intubation**<br>**suction** | 19 | **6.0** [0-11] | P < 0.0001 | P>0.99 |
| **Pre-extubation**<br>**suction** | 14 | **3.1** [0-6.5] | P <0.0001 | P>0.99 |
| **Extubation** | 14 | **11** [3.6-28] | P = 0.1 | P = 0.6 |
| **Post-extubation**<br>**suction** | 13 | **12** [0-30] | P = 0.029 | P>0.99 |

**Figure 2.**
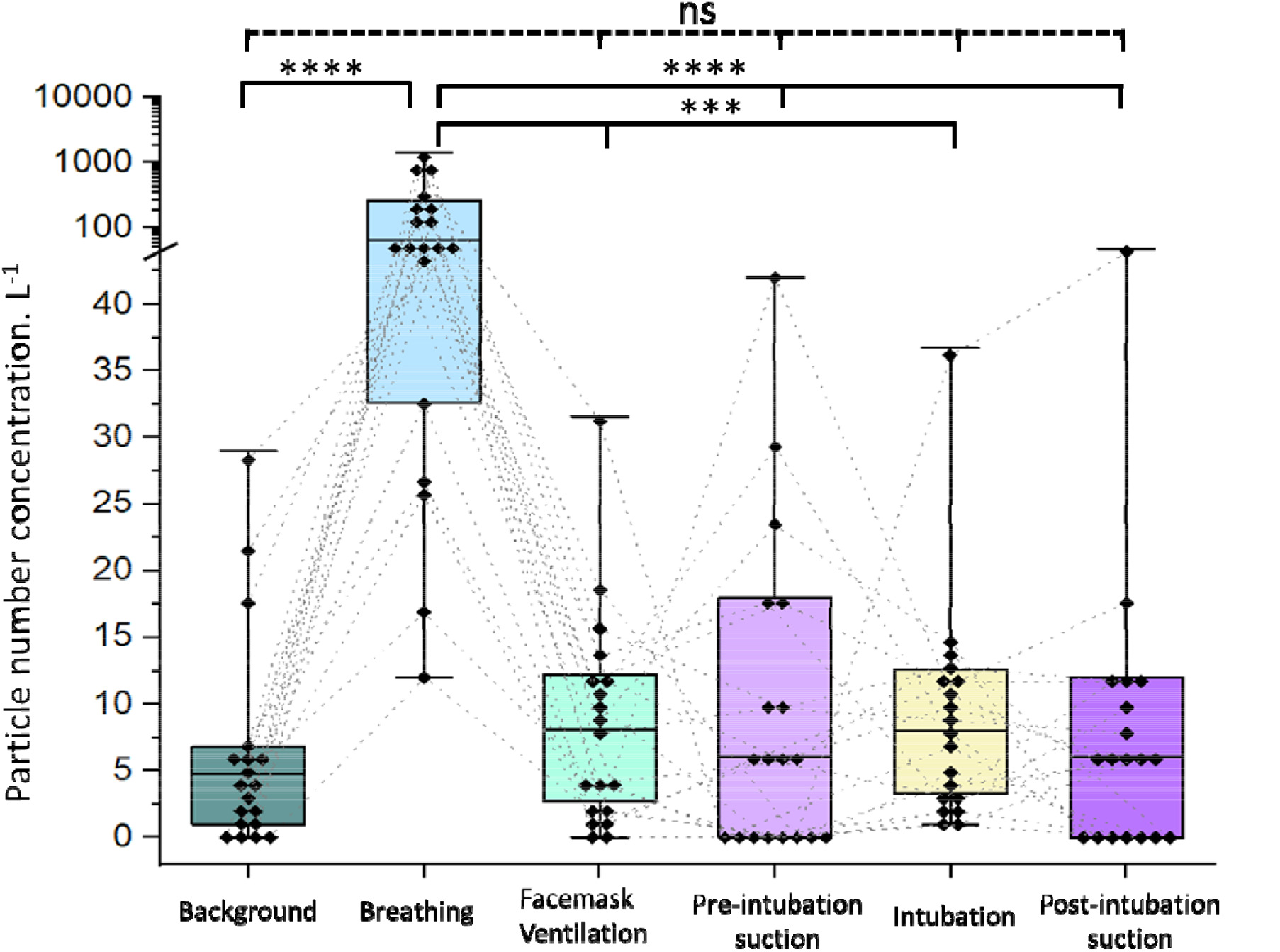
Plot of aerosol concentrations for the phases of the tracheal intubation sequence including suction compared to breathing and background aerosol concentrations. Both episodes of airway suction were associated with lower concentration of aerosol than the patients’ tidal breathing (likewise for facemask ventilation and intubation). Boxes = IQR, whiskers = range, solid horizontal line = median, • = individual patient data, dotted lines connect individual patient’s data points. (n=19 except facemask ventilation (n=18), **** = p<0.0001, *** = p<0.001, ns = non-significant, Friedman with Dunn’s multiple comparisons). Note linear - log scale representing the aerosol concentrations to cover the wide range of values seen with tidal breathing.

**Figure 3.**
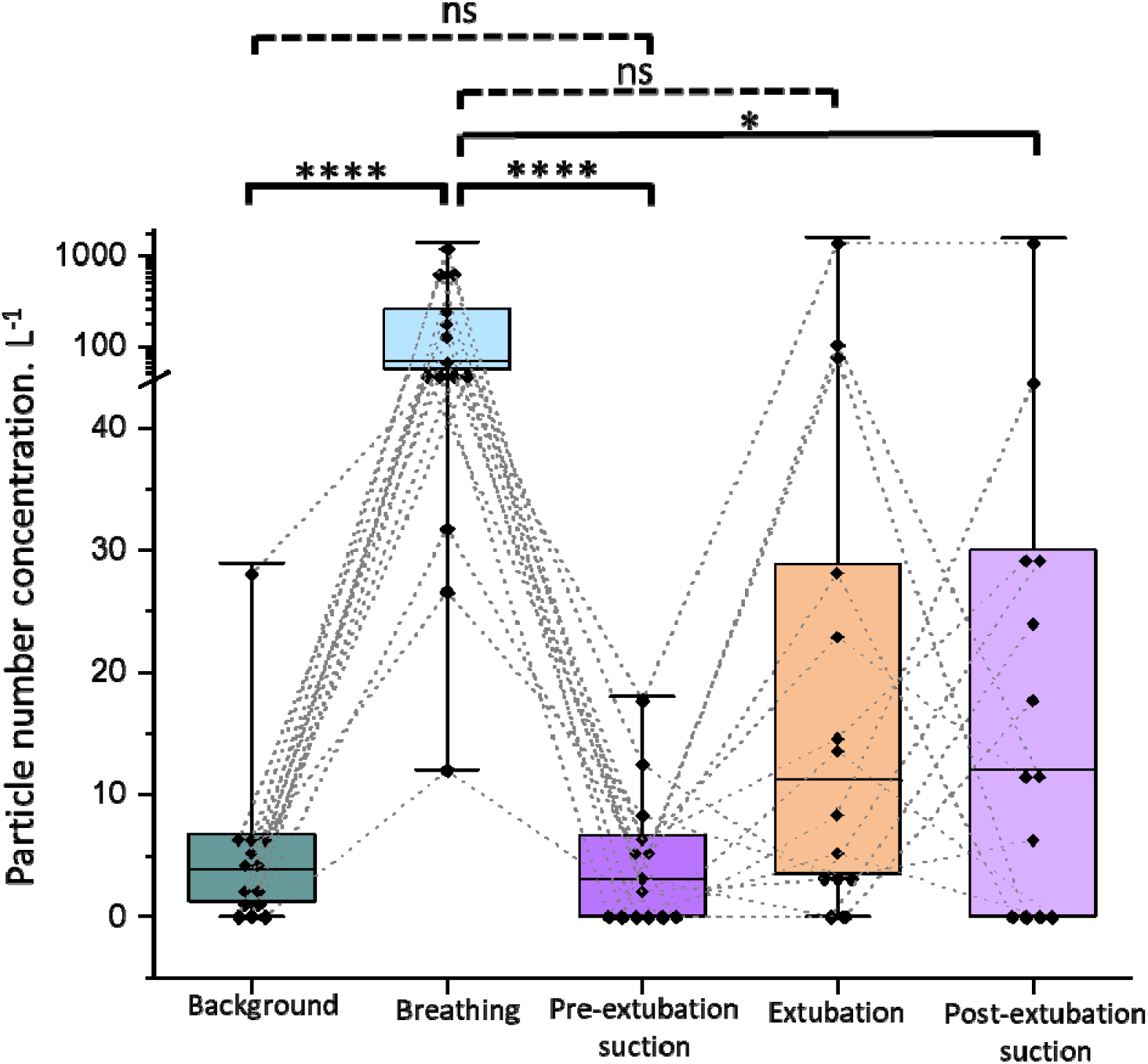
Aerosol concentrations for the phases of tracheal extubation including suction compared to tidal breathing and background. Upper airway suction with a secured (intubated) airway is associated with lower aerosol concentration than tidal breathing and is no different to background. Upper airway suction in an open airway in a spontaneously breathing patient is not significantly different to that seen with tidal breathing. Boxes = IQR, whiskers = range, solid horizontal line = median, • = individual participants data connected with dotted lines. (n=14 except post extubation suction (n=13), **** = p<0.0001, * = p<0.05, ns = non-significant, Friedman with Dunn’s multiple comparisons). Note linear - log scale representing the aerosol concentrations to cover the wide range of values seen with tidal breathing.

Aerosol concentrations recorded during extubation (11 [3.5-12] particles.L^−1^) and during post-extubation suctioning (12 [0-30] particles.L^−1^) showed a wide spread of values (Figure 3) however, extubation aerosol concentrations were not significantly different to awake tidal breathing and post-extubation suctioning was lower than for tidal breathing (p = 0.1 and p = 0.029 respectively, Friedman). The average aerosol concentration recorded during all periods of suction was 6.0 [5-30] particles.L^−1^ which was much lower than breathing (p = 0.0007, Friedman).

The peak aerosol concentrations produced by volitional coughs (1520 [600-4363] particles.L^−1^) and tidal breathing (540 [240-1680] particles.L^−1^) were many-fold higher than the peak concentrations recorded during all periods of upper airway suctioning (45 [30-90] particles.L^−1^, both p<0.0001, Friedman) (Figure 4).

**Figure 4.**
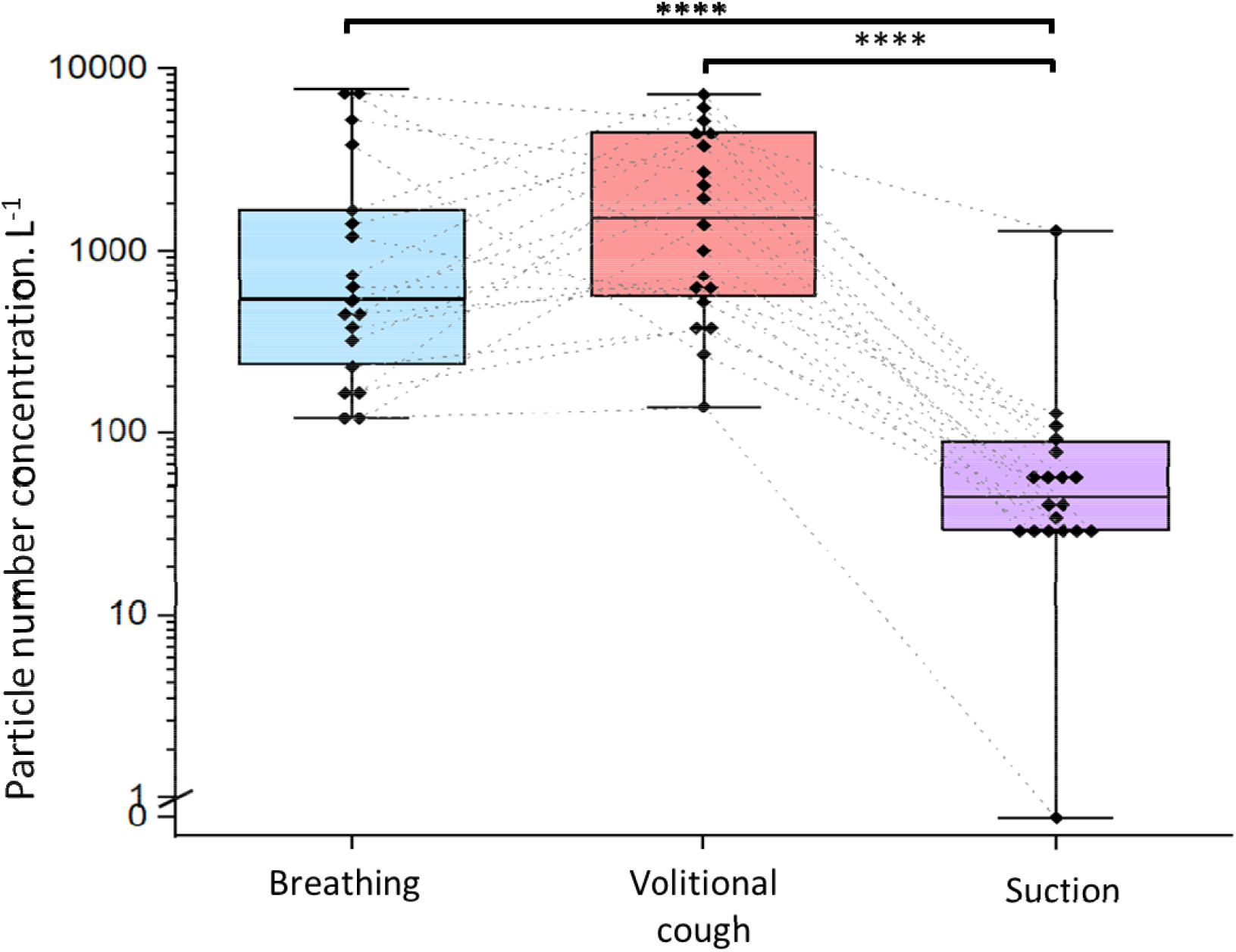
Peak aerosol concentrations during breathing and coughing compared to the peak aerosol concentration measured during all periods of airway suction. Boxes = IQR, whiskers = range, dotted lines connect individual patient’s data points, solid horizontal line = median, • = individual participants. (n=19, **** = p<0.0001, Friedman with Dunn’s multiple comparisons).

## Discussion

Managing upper airway suction as an AGP impacts theatre efficiency; when performed before tracheal extubation, all healthcare workers present in the operating theatre must either wear airborne protection PPE or leave theatre. Evacuation of team members at the end of the case prevents theatre cleaning and preparation for the next patient, until a period of ‘fallow time’ has been observed. Our study shows that upper airway suctioning in an anaesthetised patient is associated with aerosol concentrations that are indistinguishable from background, and much lower than those seen during breathing and coughing. Upper airway suctioning in the anaesthetised patient should therefore not be designated or managed as an AGP. When upper airway suction is performed following tracheal extubation, when a patient is breathing and able to cough, then the aerosol concentrations generated are no higher than those produced by natural respiratory events in the same patient. Upper airway suctioning in the awake patient does not of itself generate aerosol and adds no additional risk to the patient’s own natural respiratory activities.

The application of upper airway suction of the laryngopharynx in anaesthetised patients with a secured airway (tracheal tube *in situ*) represents the best approximation of studying the procedure alone, independent of any aerosol generation from the patient’s own respiratory activity. This is represented in this study by airway suctioning post-intubation and pre-extubation. It is reassuring that we saw no increase in the aerosol concentration during these suction procedures indicating that upper airway suctioning is not intrinsically an AGP.

We found that upper airway suctioning of the open airway in a spontaneously breathing patient (after extubation) does not increase the level of aerosol generated above that produced by the patient’s own natural respiratory activities. In this current study, one patient coughed following tracheal extubation and again during their subsequent period of suctioning which generated aerosol. Patients may cough during or following any airway manipulation. If this natural respiratory activity is of concern to the practitioner (due to a risk assessment indicating potential risk of COVID-19) then we suggest that all close patient interactions should be performed whilst wearing airborne protection PPE as the patient will be generating respiratory aerosol with every breath they exhale [9–11]. It should also be noted that placement of a facemask over a patient’s mouth and nose will prevent the escape of aerosol generated during a cough after extubation [5].

This study extends the clinical aerosol methodology developed by the AERATOR group. The strengths of the study include the use of continuous aerosol sampling during airway management with event timestamping, which enables accurate comparison of the components of airway management for each patient; within patient comparisons account for the known large variation between subjects and an ultraclean theatre environment enables resolution of respiratory aerosol from breathing and coughing above background. Our results reflect real world practice having captured airway management requiring oropharyngeal airway devices, intubation aids and management of an unintended oesophageal intubation. Additionally, one patient had hiccoughs following tracheal intubation and bit down on the tracheal tube during extubation for over 90 seconds. There was no increase in aerosol concentrations during any of these events. The procedures were performed by anaesthetists ranging from novice to consultant and as such, our findings are generalisable to a wide variety of settings.

The study has some weaknesses. It is a single centre study, and the numbers of patients are relatively small. However, the high-resolution sampling enabled clear identification of respiratory aerosol and the study’s power to identify group level differences is enhanced by the within-subject design. We are also unable to identify whether the recorded aerosol is carrying virus or make an absolute assessment regarding the risk of transmission. Of note the degree of risk associated with absolute viral levels is unknown.

There seems to be no plausible mechanism for aerosol generation by the procedure of upper airway suctioning. Suction canisters operate by applying a sub-atmospheric pressure and are designed to draw secretions into the tubing and away from the site of the suction device, and therefore from the practitioner. Of note, the use of suction in the oropharynx has been shown to reduce the amount of aerosol generated during other aerosol generating procedures such as oesophagoduodenoscopy and dental procedures [12–15]. In contrast, upper airway suctioning performed in a non-anaesthetised patient may evoke coughing which can generate high levels of aerosol. As such, the procedure is not aerosol generating but the patient can generate respiratory aerosol through natural respiratory activities.

During the conduct of the study, we recorded aerosol generation through a further 19 entire intubation sequences and 14 extubation sequences (including periods of facemask ventilation) which produced findings consistent with our previous study with low levels of aerosol generation compared to cough [6,7]. A total of 19 patients were recruited to ensure the study was adequately powered to detect differences during post extubation suctioning which was more difficult to obtain (n=13). The current study extends our previous findings by applying more stringent criteria through recording aerosol concentrations closer to the patient (at 20cm vs 50cm) and by comparing the patient’s own tidal breathing and coughs against procedural aerosol generation. In all cases this confirmed that tracheal intubation, extubation and facemask ventilation are low risk for aerosol generation. Thus, in combination with our observation that upper airway suction is not aerosol generating, we have increased confidence to assert that none of the common components of tracheal intubation and extubation sequences in anaesthetised patients should be considered as AGPs.

Our findings add further weight to the need to reassess the list of ‘Aerosol Generating Procedures’ or better even to reassess use of the term at all [11]. This term places the emphasis of risk on the medical procedure (many of which have been demonstrated not to generate aerosol [6,16-19]) whilst neglecting multiple factors that should be considered during the risk of assessment of any patient interaction. The risk assessment should consider the likelihood of the patient being infected with a respiratory pathogen (such as SARS-CoV-2), the proximity of the operator to the patient’s respiratory tract, the duration of this proximity, the health, immune and vaccination status of the operator and the environment in which the interaction occurs. The results of this study again lead us to conclude that the focus and emphasis regarding aerosolised pathogen transmission risk should be on the patient rather than specifically the procedure.

## Data Availability

All data produced in the present study are available upon reasonable request to the authors

## Acknowledgments

The AERATOR study is registered in the ISRCTN registry (ISRCT:N21447815). Andrew Shrimpton is an NIHR funded doctoral Research Fellow, the MAGPIE study is part of the NIHR301520 grant. AERATOR is funded by an NIHR-UKRI rapid rolling grant (Ref: COV0333). *This report presents independent research commissioned by the National Institute for Health Research (NIHR). The views and opinions expressed by authors in this publication are those of the authors and do not necessarily reflect those of the NHS, the NIHR, UKRI, or the Department of Health*.

## Competing Interests

AEP declares advisory board work for Lateral Pharma and consultancy for and research grants from Eli Lilly for projects unrelated to this study.

## Notes

### Author Declarations

Ethical approval was granted by the Greater Manchester REC committee (Reference: 20/NW/0393, approved 18/09/2020) and all patients gave written informed consent. The study was granted Urgent Public Health status by NIHR and is registered in the ISRCTN registry (ISRCT:N21447815).

